# Borderline personality disorder following resection of large sagittal sinus meningioma is evidenced by a self-other voice discrimination task: a case report

**DOI:** 10.1101/2022.08.28.22279288

**Authors:** Pavo Orepic, Giannina Rita Iannotti, Julien Haemmerli, Cristina Goga, Hyeong-Dong Park, Sophie Betka, Olaf Blanke, Christoph M. Michel, Guido Bondolfi, Karl Schaller

**Author notes:** **Corresponding author** Prof. Karl Schaller, Department of Neurosurgery, Geneva University Medical Center & Faculty of Medicine, University of Geneva, Switzerland, Hôpitaux Universitaires de Genève, Rue Gabrielle-Perret-Gentil 4, 1205 Geneva, Switzerland. The two authors contributed equally to the conception of study, analyses and writing of the manuscript (Pavo Orepic, Giannina Rita Iannotti).

## Abstract

Personality changes following neurosurgical procedures pose a major concern for patients and remain poorly understood both by clinicians and neuroscientists. Here we report a case of a female patient in her 50s who underwent resection of a large sagittal sinus meningioma with bilateral extension, including resection and ligation of the superior sagittal sinus, that resulted in borderline personality disorder and symptoms resembling the Gastaut-Geschwind syndrome. Clinical observations were further reflected and experimentally quantified with a series of behavioral and neuroimaging tasks assessing self-other voice discrimination, one of the established markers for self-consciousness. In all tasks, the patient consistently confused self- and other voices – i.e., she misattributed other-voice stimuli to herself and self-voice stimuli to others. Moreover, behavioral findings were corroborated with scalp EEG results. Specifically, the same EEG microstate, that was in healthy participants associated with hearing their own voice, in this patient occurred more often for other-voice stimuli. We hypothesize that the patient’s preexisting psychological problems were significantly aggravated by postoperative decompensation of a fragile steady-state combination of direct frontal lobe compression and preoperative development of a large venous collateral hemodynamic network that followed gradual occlusion of the superior sagittal sinus. Resection of the sagittal sinus together with the tumor impacted venous drainage of brain areas associated with self-consciousness. These findings are of high relevance for developing experimental biomarkers of post-surgical personality alterations.

## INTRODUCTION

Fear of post-operative changes in personality and the sense of self remains a major source of distress for patients undergoing a neurosurgical procedure (Schaller et al., 2021). Neurosurgeons are thus placed in a precarious situation, being unable to provide feedback about the likelihood of such outcomes, as both their frequency and circumstances remain largely unknown. Hence, there is a strong need for documented reports addressing post-surgical personality changes, in order to gain sufficient knowledge potentially leading towards established protocols and clinical evaluations related to post-surgical alterations of the sense of self.

Sense of self has become one the key interests in recent neuroscience research, potentially opening the door to the expertise necessary to bridge the aforementioned clinical gap. Namely, there has been an increase of experimental paradigms developed to assess various aspects of self-consciousness. Broadly, such paradigms could be divided in two categories (Gallagher, 2000) – those related to the bodily aspects of the self (Blanke, 2012; Blanke et al., 2015; Park & Blanke, 2019), such as multisensory and sensorimotor integration, and those related to cognitive aspects (Feinberg & Keenan, 2005; Northoff et al., 2006; Qin et al., 2020), such as language, memory, or recognition of self-related cues. Combining those paradigms with neuroimaging enables to pinpoint brain areas that scaffold the sense of self, thereby serving as potential candidates related to post-operative personality alterations. They often include insular, medial prefrontal, cingulate, medial prefrontal, and inferior frontal cortices, as well as temporo-parietal junction (Blanke et al., 2015; Legrand & Ruby, 2009; Northoff et al., 2006; Park & Blanke, 2019; Scalabrini et al., 2021). Compared to healthy controls, patients often perform differently in experimental paradigms assessing the sense of self (Bassolino et al., 2019; Bernasconi et al., 2021; Betka et al., 2022; Candini et al., 2018; Schaller et al., 2021; Shergill et al., 2005; Whitford, 2019), which is indicative of a blurred self-other boundary in certain pathologies. For instance, inability to distinguish self from other has long been related to some psychiatric symptoms, such as passivity sensations and auditory-verbal hallucinations (i.e. ‘hearing voices’) (Ford & Mathalon, 2005; Frith, 1987; Frith et al., 2000; Moseley et al., 2013; Shergill et al., 2005; Whitford, 2019).

One such paradigm is our self-other voice discrimination (SOVD) task (Orepic et al., 2021; Orepic, Kannape, et al., 2022; Orepic, Park, et al., 2022), that belongs to the second category and enables to draw a perceptual boundary between self and other in the auditory domain. With our SOVD task, we are able to pinpoint individual perceptual specificities related to auditory self-other boundary (Orepic, Kannape, et al., 2022; Orepic, Park, et al., 2022), such as impairments in general self-other discriminability, inability to recognize only self/other voice, or a bias to hear self/other voice. Moreover, we identified a self-voice specific EEG microstate, at mean latency 345 ms, during SOVD in healthy population (Iannotti et al., 2021) that activates a brain network involving insula, cingulate cortex, and medial temporal lobe structures. This microstate occurs more often when participants hear self-voice, compared to hearing other voices, and this correlates with SOVD task performance. Thus, assessing SOVD and the corresponding neural activity in patients might serve as a biomarker of pathological alterations in the sense of self, including post-surgical personality alterations.

Here, we report a case of a patient who experienced a significant personality change that was significantly intensified following a neurosurgical procedure, and eventually resulted in a diagnosis of borderline personality disorder. Importantly, performance in the aforementioned SOVD task was concordant with her neuropsychological and psychiatric reports. Specifically, the patient reported hearing her own voice when, in fact, she was presented with the voices of others, and vice versa – she reported hearing her voice when presented with other-voice stimuli. Remarkably, the behavior opposite to healthy participants was even corroborated in task-related EEG findings – the neural pattern observed in healthy participants associated with self-voice (Iannotti et al., 2021), was observed in this patient with other-voice stimuli. In the sections below, we summarize her clinical image and describe behavioral as well as neuroimaging analyses and results.

## NEUROSURGICAL PROCEDURES

In 2014, the patient underwent a bi-frontoparietal craniotomy for the resection of a large superior sagittal sinus meningioma with bilateral extension, including resection and ligation of the superior sagittal sinus. Preoperative MRI showed a left frontal 5 × 4 cm extra-axial lesion invading the anterior portion of the superior sagittal sinus (Figure 1A). An electroencephalogram was performed prior to the surgery and did not reveal any pathological findings. The surgical removal of the lesion Simpson IV underwent without any complications. No new focal neurological deficit could be observed at the postoperative time. The final diagnostic revealed a transitional meningioma WHO I. The postoperative MRI showed a small residual tumoral tissue at the posterior superior portion of the resection cavity (Figure 1B). The yearly MRI showed no recurrence of the meningioma and a stability of the small remanent until five years after surgery, in which the remanent appeared larger (Figure 1C). A second surgery was performed in 2020, and the recurrent meningioma could be removed without complication. No new focal neurological deficit was observed.

**Figure 1:**
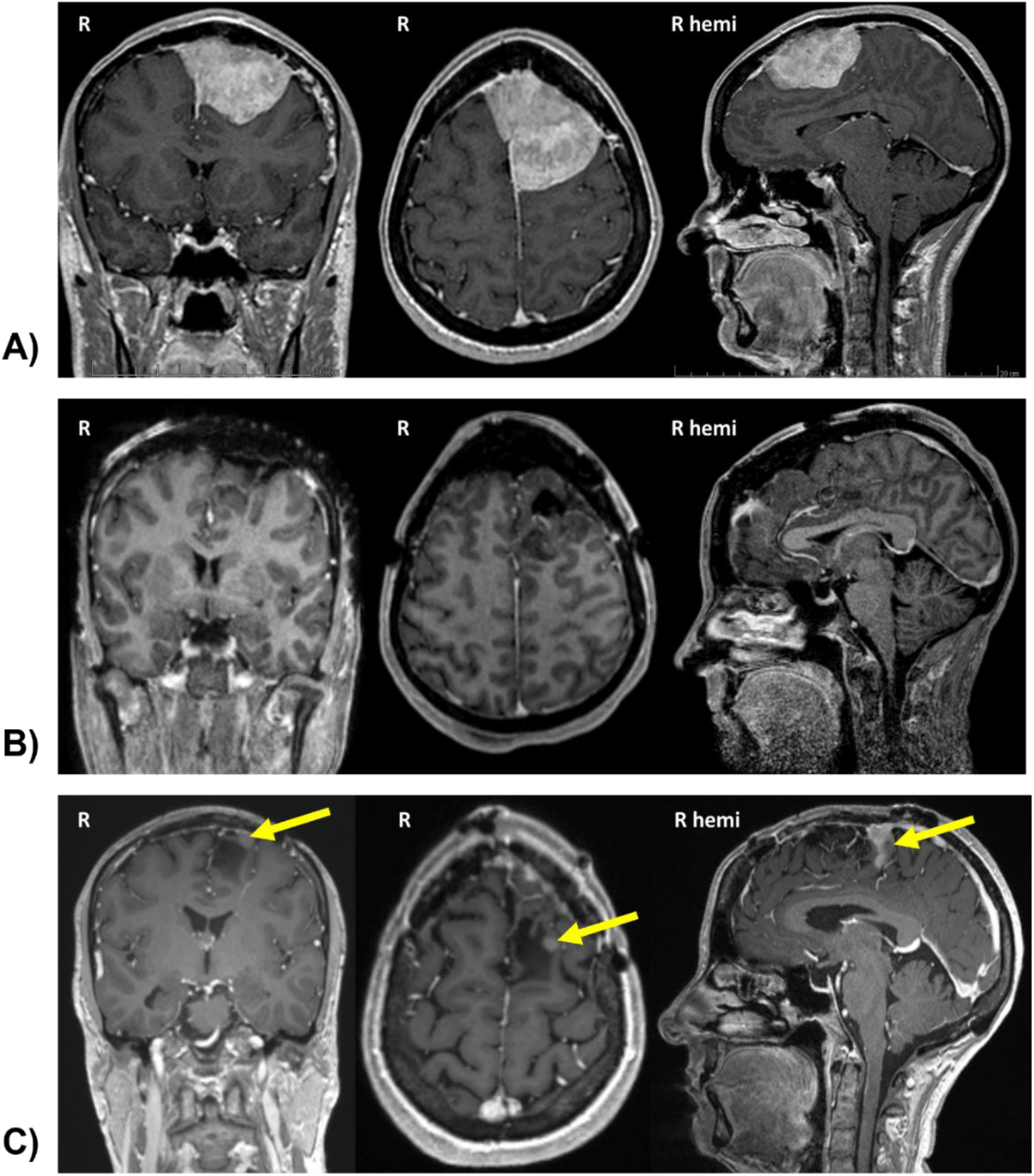
Patient’s structural imaging. **A)** The preoperative structural 3D-T1 with Gadolinium revealed a left frontal extra-axial lesion (meningioma) extended to the anterior sagittal sinus. **B)** MRI following the neurosurgery, a left parasagittal craniotomy, showed a successful removal of the tumor **C)** A subsequent MRI (1-year follow-up) indicated a residual of the meningioma (yellow arrow).

## NEUROPSYCHOLOGICAL AND PSYCHIATRIC EVALUATIONS

### Before meningioma resection

In 2012, two years before the removal of the meningioma, the patient started complaining about “*feeling different*”. She described experiencing changes in “*personality*” and “*behavior*”, without being able to specify the causes of those changes. Clinical investigations identified polymorphic psychiatric symptomatology characterized by atypical anorexic behaviors, emotional dysregulation with episodic self-damaging behaviors, panic attacks, and periodical alcohol abuse. She also reported sensory disturbances in the form of paraesthesias along her right leg and dysesthesias along her right index finger that would disappear after 24 hours. Physical exams did not show any pathological signs, in particular no sensory deficits. The neuropsychological exam prior to surgery did not report anything pathological, apart from an increase in anxiety, likely due to the proximity of the surgery.

### After meningioma resection

After the removal of meningioma (in 2014), the aforementioned personality alterations decompensated. Mood fluctuations were more frequent and more intense, the quality of sleep degraded, anxiety levels were higher, and depressive traits appeared, accompanied with suicidal thoughts. The patient complained about “*feeling of no longer being able to understand herself*”, and about “*the fear of losing control over her body*”. She reported “*becoming more and more harsh towards herself while at the same time feeling softer and more tolerant towards other*s”. The neurophysiological exam following the surgery was still unable to highlight anything particularly pathological, apart from an increase in depressive traits and a possibility for ADHD.

As time went by, her psychological disturbances worsened and emotional liability increased. The patient had two suicide attempts and two isolated psychotic episodes with complex and non-malevolent multisensory hallucinations (visual, auditory, and olfactory), accompanied by feelings of presence. As a result, in 2016 (two years after the surgery) she was hospitalized for a suspicion of epileptic phenomena that could have occurred as a result of meningioma resection and partially account for the present symptoms. At the same time, psychiatric evaluation reported an ICD-10 diagnosis of organic emotionally labile (asthenic) disorder (F06.6) and emotionally unstable personality disorder, borderline type (F60.31). F06.6 related the diagnosis to the lesional effect of the meningioma. Following the meningioma resection, the patient also developed symptoms resembling the Gastaut-Geschwind syndrome – a hypergraphia, marked hyposexuality, and an increased interest in subjects related to spirituality, that was absent throughout his previous life, but without particular religiosity. A neurological examination from 2017 reported a temporal epilepsy manifested with uncinate seizures, that could partially account for these symptoms (Devinsky & Schachter, 2009; Tebartz van Elst et al., 2003). Approximately a year after the neurological report, in 2018, the patient participated in our experiment. The chronological timeline of the described events is illustrated on Figure 2.

**Figure 2:**
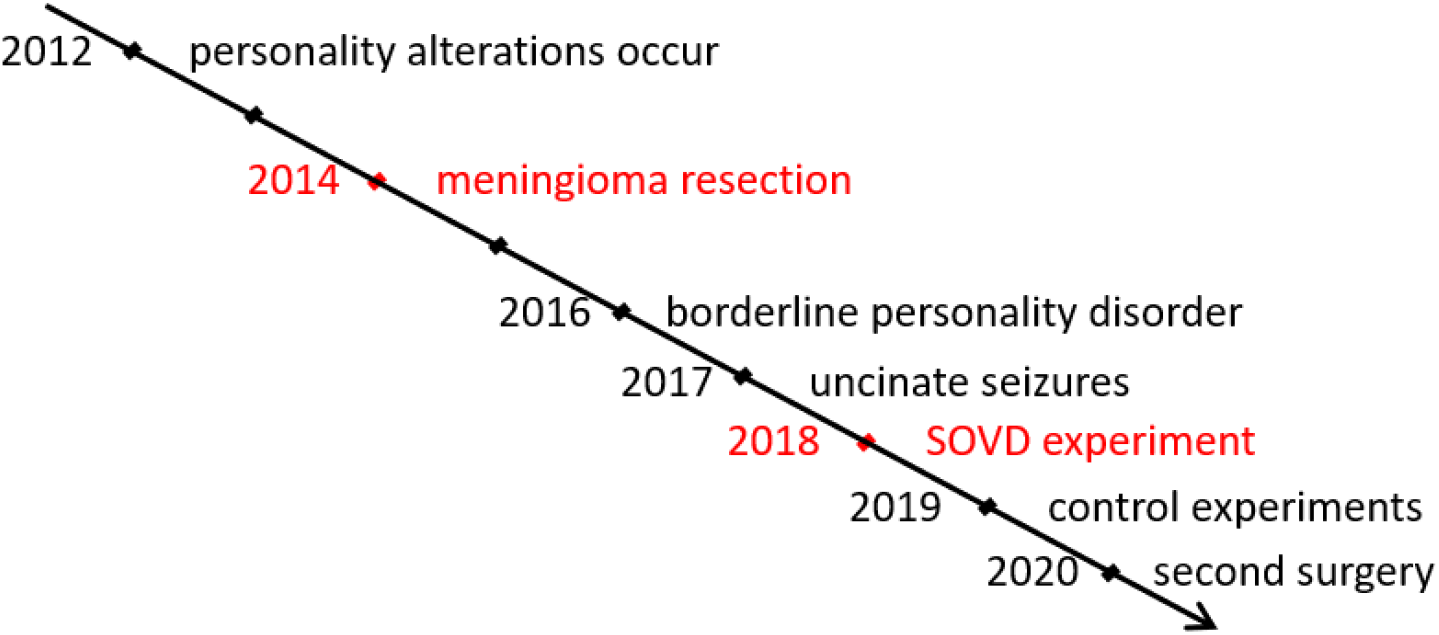
Timeline outlining patient’s medical history.

## BEHAVIOURAL TASKS AND RESULTS

The current patient (further referred to as ‘patient 4’) (patient IDs are not known to anyone outside the research group) belonged to a cohort of 24 patients that were recruited to assess and quantify the effects of surgical resection of various brain regions on potential changes in self-consciousness with carefully designed experimental paradigms. One such paradigm is our self-other voice discrimination task (Orepic, Kannape, et al., 2022; Orepic, Park, et al., 2022), in which participants try to identify the dominant voice (self or other) in ambiguous self-other voice morphs, allowing us to quantify the perceptual self-other boundary in the auditory domain. Specifically, patient’s voice (vocalization /a/ with the duration of 500 ms) was morphed with a voice of a gender-matched unfamiliar voice, generating vocal stimuli that contained different ratios of patient’s voice (e.g., a voice morph could contain 40% of patient’s voice and 60% of an unfamiliar voice). The patient heard 6 different voice morphs (containing 15%, 30%, 45%, 55%, 70%, and 85% of her voice), each repeated 50 times in a random order (total of 300 trials), and was instructed to indicate, by clicking on a button, whether the voice she heard more resembled her voice or the voice of someone else. Responses to this task enable fitting of psychometric curves, reflecting the characteristics of individual performances in self-other voice discrimination task – e.g., a steeper curve indicates an overall increase in performance, a rightwards or leftwards shift of the curve reflect a bias to hear other or self-voice, respectively, etc. To decrease task difficulty, the unmorphed voice was once presented to the patient prior to the task. For a discussion on the task parameters and their effect on the behavior of healthy participants see (Orepic, Kannape, et al., 2022).

Cyan psychometric curve on Figure 3 indicates average behavior of all other patients (individual psychometric curves and their relationship to the respective resections will be reported elsewhere), indicating a typical increase in ‘self’ responses (y axis) with the increase of self-voice ratio present in voice morphs (x axis). The red psychometric curve on Figure 3, that of patient 4, has a negative slope, indicating that the patient inverted self- and other voices (i.e., she was responding ‘self’ for other-dominant voice morphs, and vice versa). Absolute value of the slope of the patient’s curve was very similar to the average slope of other patients (patient 4: 83.4%, other patients: mean=80.6%, SD=6.9%), indicating that the patient could indeed discriminate between the two voices, but despite good discriminability, misattributed other-voice stimuli to herself and self-voice stimuli to someone else.

**Figure 3:**
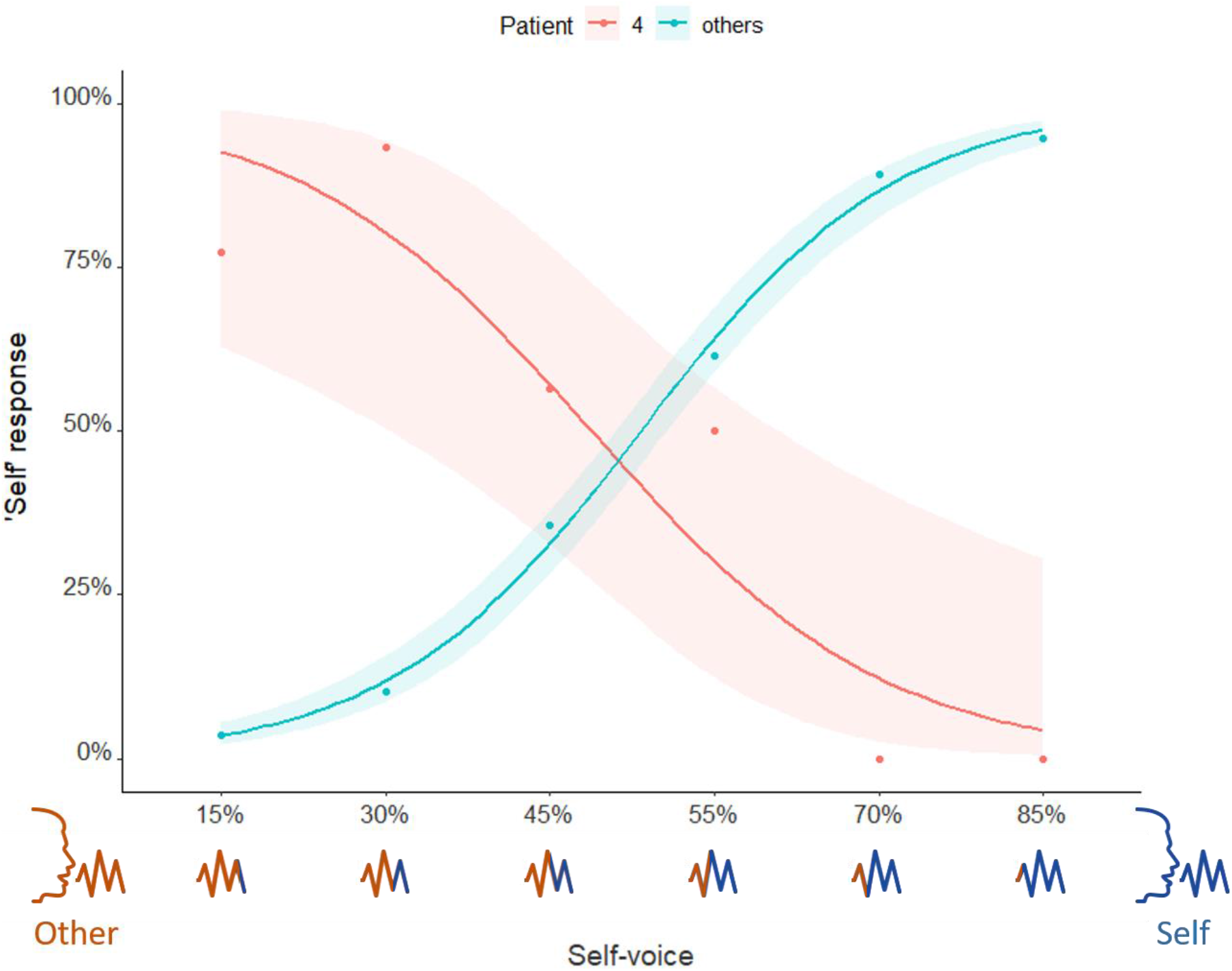
Psychometric curves fitted for patients’ performance in self-other voice discrimination (SOVD) task. The abscissa indicates the percentage of the self-voice present in a voice morph and the ordinate indicates the rate at which the corresponding voice morph was perceived as resembling the self-voice. The shaded areas around each curve represent the 95% confidence intervals. Contrary to all other patients (cyan), patient 4 (red) inverted self- and other voices.

In order to investigate whether the observed self-other confusion was specific to this task, on a second visit (in 2019), we performed three additional control tasks that involved no voice morphing (i.e., 100% or 0% self-voice). In all tasks, the same /a/ vocalizations were used in total of 20 counterbalanced trials, presented in a randomized order. In the first task (Figure 4A), the patient heard both unmorphed voices (self and other) one after another, and was instructed to indicate which of the two voices was her voice, by pressing a button. In all 20 trials, the patient inverted the voices (i.e., responded ‘self’ for other-voice stimuli, and vice versa). In the second task (Figure 4B), the patient heard one voice per trial (self or other) and was instructed to indicate whether it was her voice. Again, in all 20 trials, the patient inverted the 2 voices. Finally, in the third task (Figure 4C), the patient was instructed to say /a/ for approximately one second, after which one voice (self or other) was immediately presented to her. When the stimuli were presented directly after speaking, the inversion rate was reduced – in 16 out of 20 trials (80%). Interestingly, in all the trials in the third task in which the patient was correct, she heard self-voice stimuli, indicating that speaking prior to the stimulus presentation increased accuracy only for the actual own voice, and not misattributed other voice.

**Figure 4:**
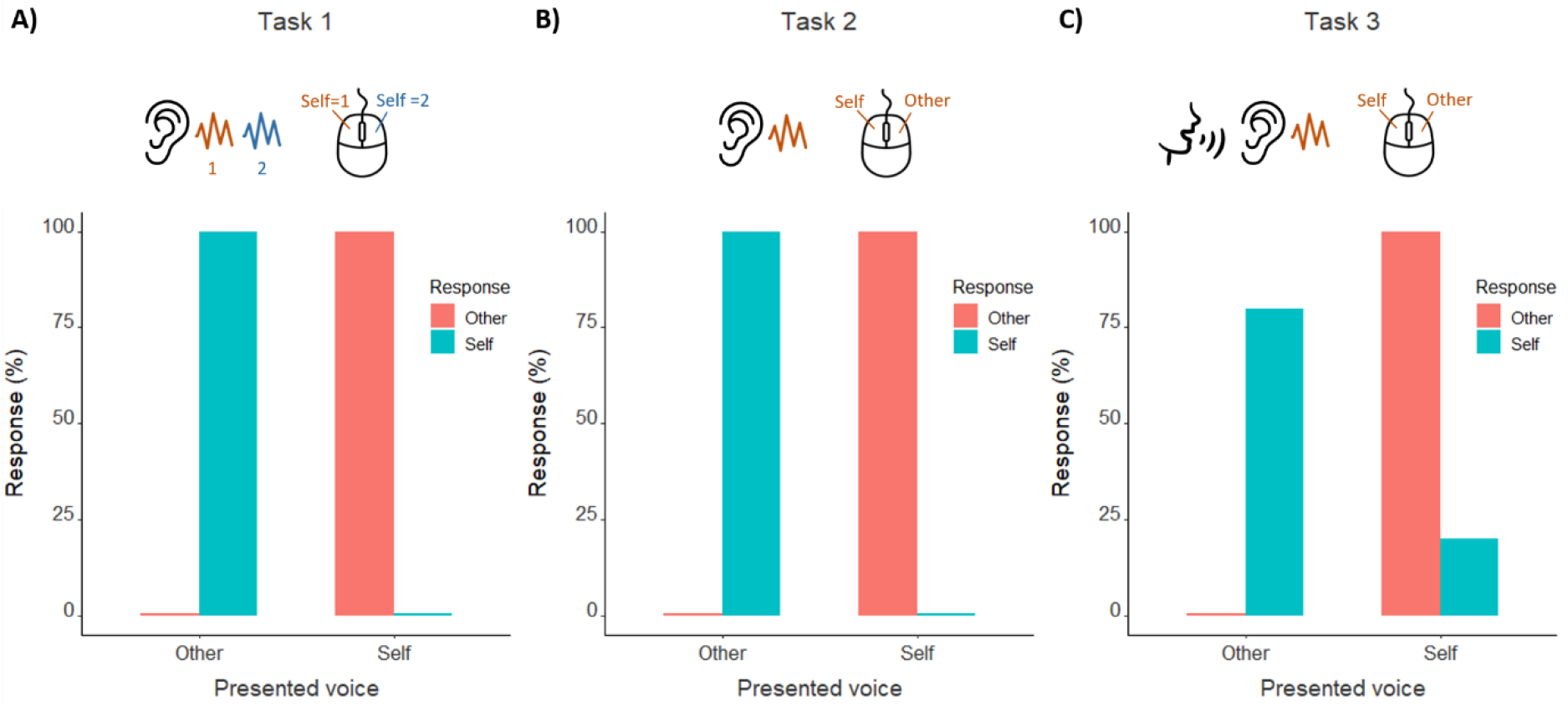
Control behavioral tasks. **A)** In task 1, after hearing two voices, the patient indicated which one corresponded to her voice. **B)** In task 2, after hearing one voice, the patient indicated whether it corresponded to her voice or someone else’s. **C)** Task 3 was the same as task 2, except that before hearing a voice, the patient produced the same utterance. In the first 2 tasks, the patient completely inverted the two voices. In task 3, a few self-voice trials were correctly recognized.

## EEG ANALYSIS AND RESULTS

During the first experimental session (in 2018), high density EEG was recorded during the self-other voice discrimination task in the setup equivalent to our previous work on healthy participants (Iannotti et al., 2021).

Event-related potentials (ERPs) analysis was conducted after EEG preprocessing, including filtering between [1-40 Hz], removal of artifacts (eye-blinking, saccades, ballistocardiogram, motion), reduction to 204 electrodes and interpolation of bad channels. ERPs were defined for each voice morph, by selecting EEG epochs between-50 and 500 milliseconds around the stimulus onset. In order to increase the number of the epochs and the signal-to-noise ratio, we grouped the ERPs belonging to each end of the self-other voice continuum. Specifically, we averaged Self-dominant (containing 85% and 70% self-voice) and Other-dominant voice morphs (15% and 30% self-voice).

EEG microstate segmentation (Brunet et al., 2011) was performed by considering the Other- and Self-dominant ERPs. This process allowed to delineate the sequence of scalp EEG topographies or microstates reflecting the brain states that are functionally stable in time (Figure 5).

**Figure 5:**
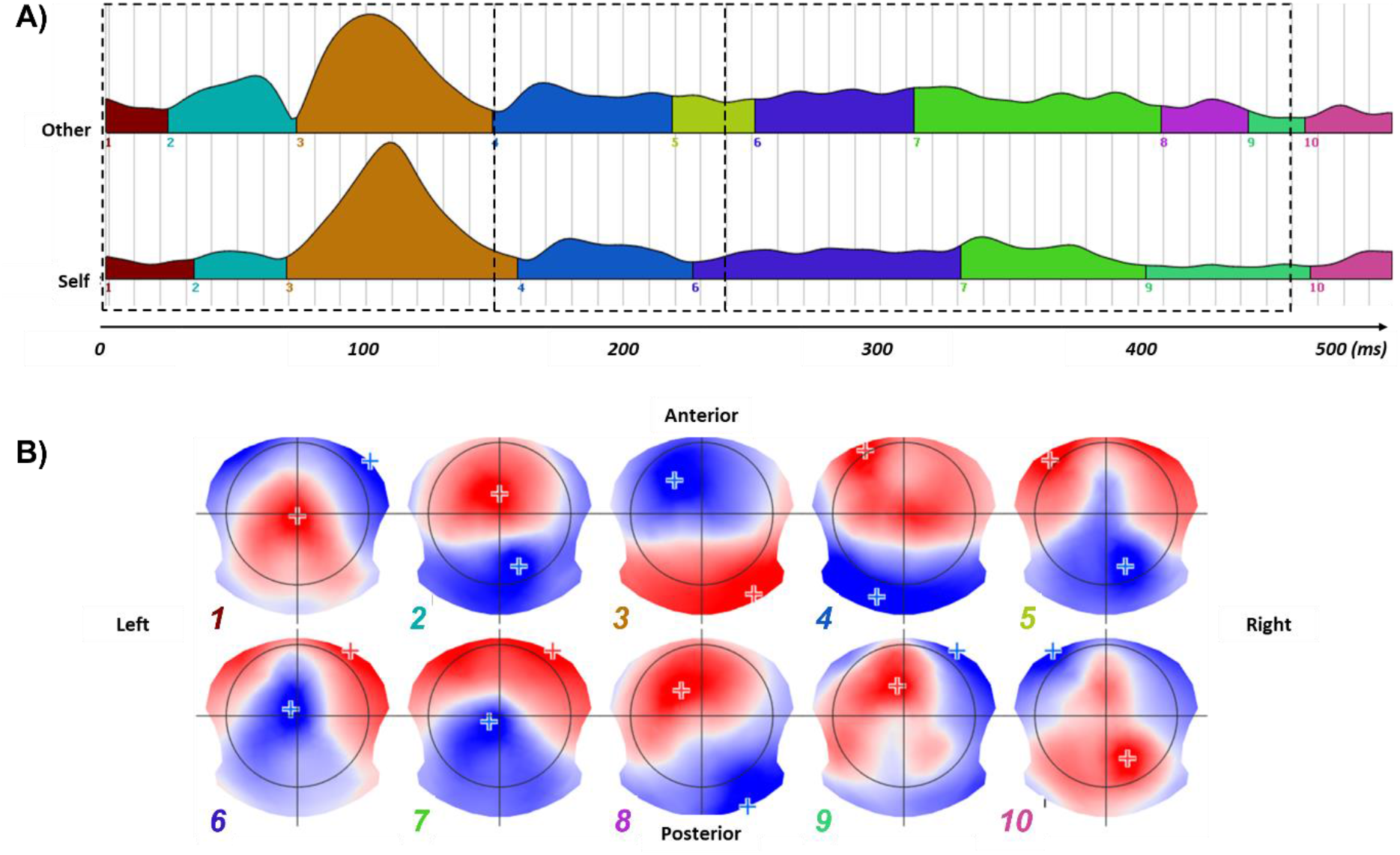
Results of the ERPs segmentation. **A)** Different colors indicate the temporal stable EEG segments resulting from the group segmentation of the Other voice (top) and Self voice (bottom). The dashed boxes indicate the three time-windows considered for the back-fitting procedure. **B)** Topographic microstates associated with each segment. Numerical labels are color-coded to match the corresponding segments. The ‘+’ symbol indicates the position of the electrodes exhibiting the highest (positive, red) and the lower (negative, blue) amplitudes of the scalp voltage potential.

To assess the statistical difference between Other- and Self-dominant morphs, we ‘fitted back’ each obtained microstate (Figure 5B) across the single ERPs (Figure 5A) associated with the two experimental conditions. Substantially, the fitting procedure assesses the spatial correlation between each microstate and the ERPs, by labelling each ERP time-point with the microstate showing the highest correlation value. In output, the fitting procedure gives the values of a set of parameters describing, among others, the power and time characteristics of each microstate along the ERPs. For each of the fitting parameters and each microstate, a paired t-test was therefore used to define the statistically significant difference between Self- and Other-dominant voice morphs. It revealed a significant difference (p<0.05) for the occurrence of microstate 7 (Figure 6A). Specifically, the occurrence of microstate 7 was significantly higher (t(96) = 2.31, p = .023) in Other-(M=60.06, SD=39.94) compared to Self-dominant (M=47.9, SD=38.96) voice morphs (Figure 6C).

**Figure 6:**
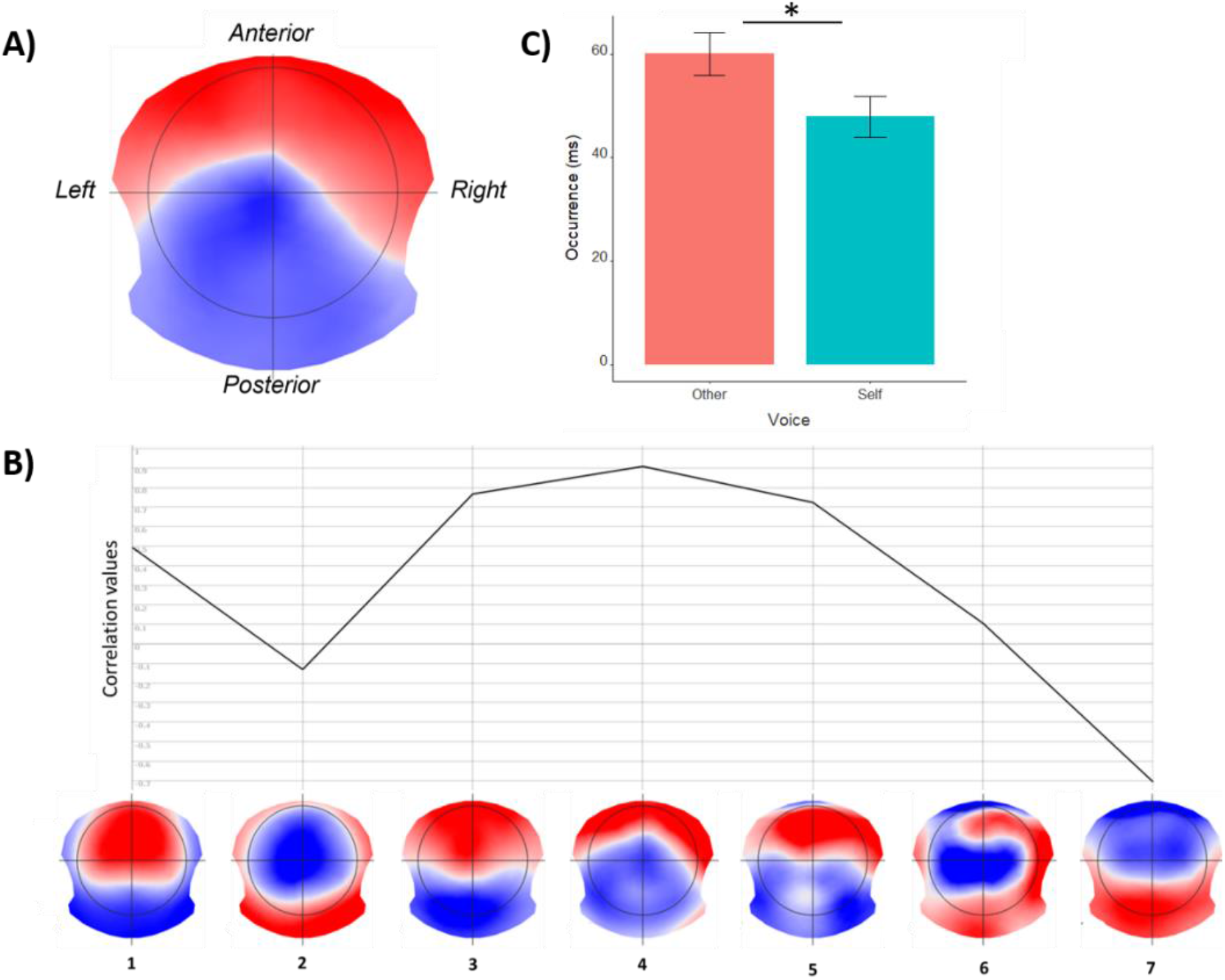
**A)** Patient’s microstate specific for the SOVD task (microstate 7 from Figure 5). **B)** Spatial correlation between patient’s microstate 7 (A) and each of the microstates observed in healthy participants while performing the same SOVD task (Iannotti et al., 2021). Patient’s microstate 7 (A) was maximally correlated with healthy participants’ microstate 4, that was SOVD-specific and occurred more often for self-voice, compared to other voice (Iannotti et al., 2021). C) Microstate 7 occurrence. Contrary to the equivalent SOVD microstate for healthy participants, patient’s microstate 7 (A) occurred more often for other voice. Bar plots indicate mean occurrence for each voice and whiskers indicate standard error.

In order to link patient’ s EEG results to our previous EEG findings on healthy participants during the equivalent SOVD task (Iannotti et al., 2021), we evaluated the spatial similarity (i.e., spatial correlation of the scalp voltage potentials) between the set of patient’s microstates and the set of healthy participants’ microstates. The highest spatial correlation (0.9, Figure 6B) was observed between patient’s microstate 7 and healthy participants’ microstate 4. Importantly, for patient’s microstate 7, an inverted pattern in respect to healthy subjects’ microstate 4 was observed. Specifically, the occurrence of microstate 4 in healthy participants was significantly higher (p<0.05) in Self-dominant compared to Other-dominant voice morphs (Iannotti et al., 2021) while the opposite was observed for patient’s microstate 7 (Figure 6C).

Finally, the neuronal sources associated with patient’s microstates were estimated by using the same electrical source imaging (ESI) approach of our previous work (Iannotti et al., 2021), by considering the patient’s post-operative MRI. We restricted the visualization to the top 10th percentile of the normalized distribution (between 0,1) of activation values across all brain. The ESI revealed that patient’s microstate 7 was associated with right-hemispheric lateralization, contralateral to the resected lesion. Two major hubs were located in the frontal and temporal lobe (Figure 7), with maximal activation in the right Inferior frontal gyrus (rIFG), opercular part.

**Figure 7:**
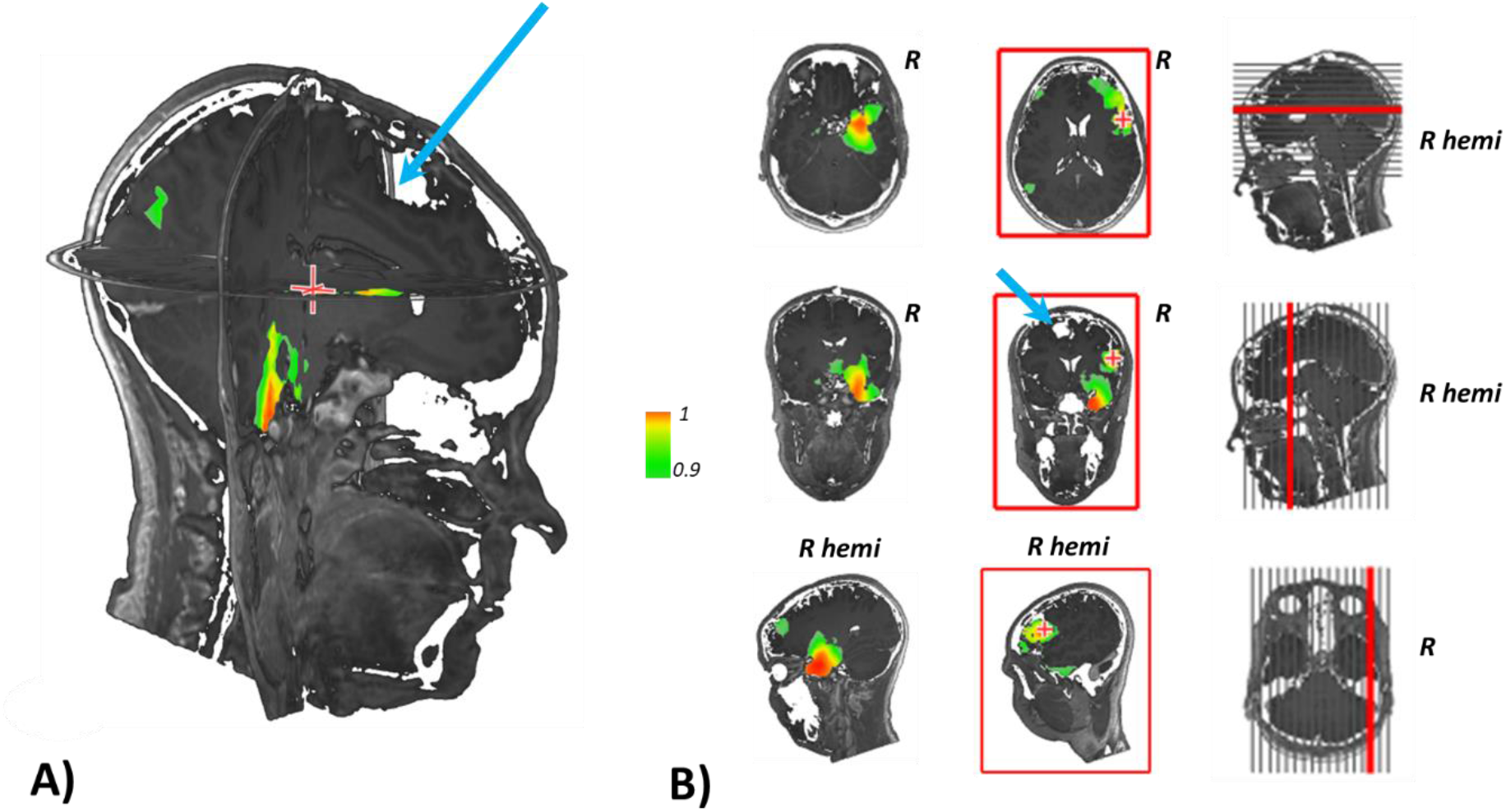
Neuronal sources associated with microstate 7. 3D **(A)** and 2D (**B)** (top to bottom: axial, coronal, sagittal) ESI results of patient’s microstate 7. Microstate 7 resulted from the concomitant activation of frontal and temporal hubs located in the right hemisphere, contralateral to the surgical resection (blue arrow in A). The visualization of activation values is restricted to the 10th percentile of the normalized ([0,1]) distribution of all activations across the brain (threshold > 0.9).

## DISCUSSION

Harmoniously joining expertise from neurosurgery, psychiatry, and neuroscience, here we report personality alterations following a neurosurgical procedure that were reflected in a sensitive neuroscientific paradigm. Recurrent feelings of detachment from patient’s mental processes that occurred following a sagittal sinus meningioma resection were reflected both in patient’s behavioral performance as well as in EEG patterns during a SOVD task.

Although personality alterations have been previously associated with frontal meningiomas (Abel et al., 2016), we hypothesize that, in our patient, they result from a combination of preexisting borderline disorder and the compressive and venous occlusive effect of the meningioma, i.e. the hemodynamic consequences associated with the meningioma (cerebral veins don’t contain valves). Hence, following gradual occlusion of the superior sagittal sinus by the meningioma, a large venous collateral network has developed, affecting the direction and the pathways of venous drainage (Brito et al., 2019). In our patient, that has concerned both frontal lobes due to the anterior sagittal sinus infiltration (Mantovani et al., 2014; Raza et al., 2010). With resection of the sinus and possible compromise of some collateral pathways and/or change of venous flow-direction, regional venous overload or alteration of regional blood flow may have impacted personality-related brain regions (Figure 8). Gradual occlusion and subsequent resection of anterior sagittal sinus could thus account for the gradual appearance of personality alterations prior to surgery, that were significantly increased following the resection.

**Figure 8:**
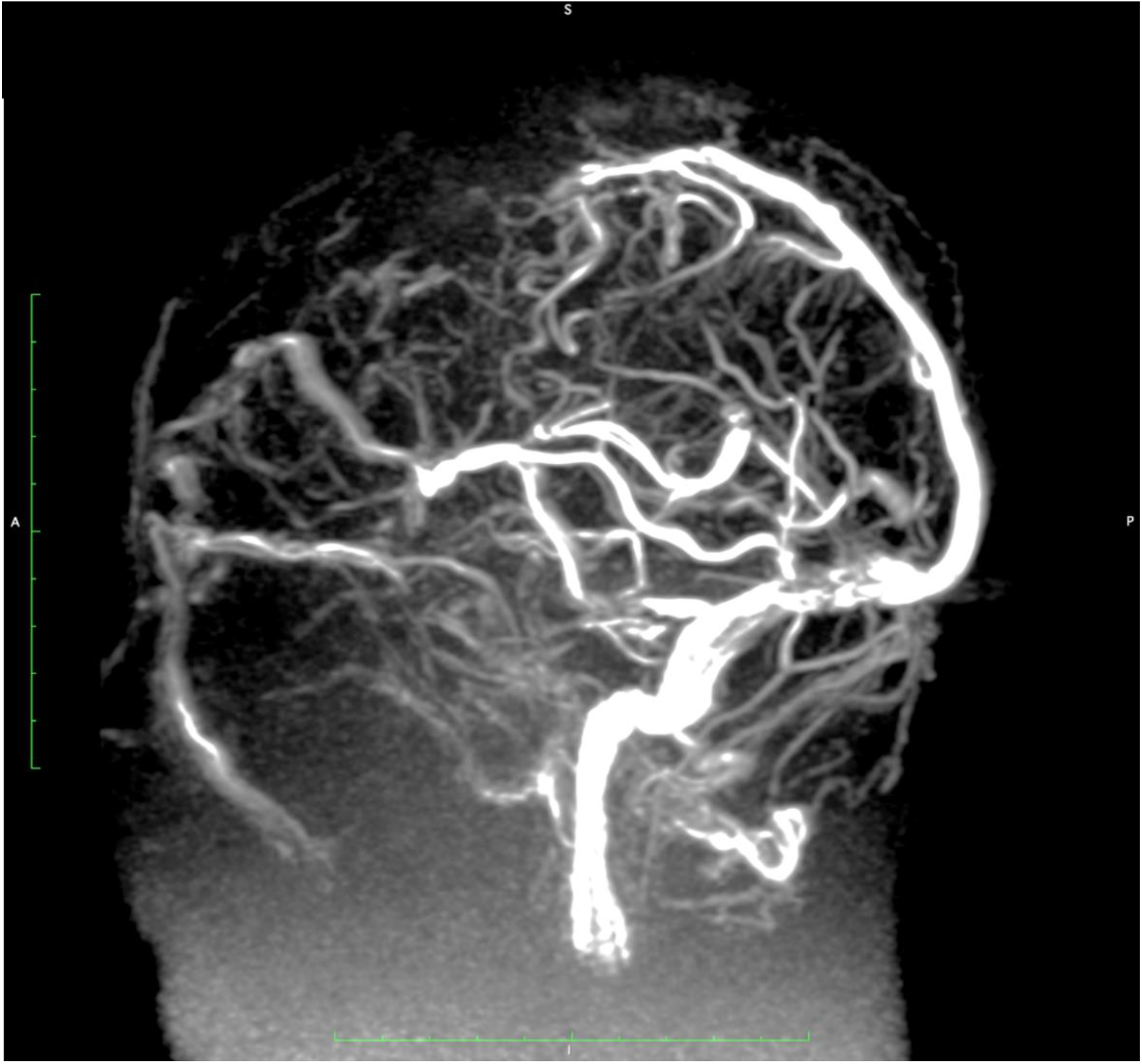
Preoperative MR venography of the patient, lateral view. The anterior part of the superior sagittal sinus is occluded. There are extensive venous collaterals anterior and posterior to the occlusion site.

Though, from a purely neurosurgical point of view that risky surgery went well, with no new postoperative neurological deficits noted, a more generalized or remote effect may be postulated. That concept is further corroborated by the delayed onset of temporal lobe epilepsy in this patient. The preoperative sensible interplay between a personality-relevant psychiatric disposition (borderline personality) and meningioma-related structural alteration of the frontal lobes and cerebral venous drainage decompensated through the open resection of the tumor and the infiltrated venous sinus.

In addition to the intellectual attempt to relating a neurosurgical action with a specific psychiatric consequence, these findings have important implications for the understanding of the brain mechanisms underlying the sense of self, which cannot be faked by the examined person in a systematic manner. First, our data relate a low-level perceptual feature – self-other voice confusion – to a higher-level deficit – borderline personality disorder, indicating phenomenological interrelatedness between different hierarchical aspects associated with the sense of the self (Blanke et al., 2015; Canzoneri et al., 2016; Northoff et al., 2006). Second, observing that brain areas associated with self-voice and the sense of self more generally (e.g., right insula (Iannotti et al., 2021; Ronchi et al., 2015; Scalabrini et al., 2021)) were in this patient associated with other-voice stimuli suggests that they might not necessarily encode the actual self-related stimuli (e.g., physical self-voice), but our subjective attribution of those stimuli. For instance, two neuroimaging studies (Kaplan et al., 2008; Nakamura et al., 2001) associated rIFG activity with hearing a physical self-voice while, in our patient, rIFG activity was associated with a voice subjectively attributed to the self, that physically belonged to another person. Potentially, one might assume that in this and related pathologies, brain activations traditionally associated with the self might be observed for any type of stimuli (e.g., faces or body parts) subjectively attributed to the self. This approach could be extended to psychiatric phenomena other than personality disorders. For instance, it has long been proposed that auditory-verbal hallucinations (AVH) – hearing voices with speakers absent – might arise as a deficit in SOVD, where internal self-vocalizations are misattributed to someone else (Ford & Mathalon, 2005; Frith, 1987; Frith et al., 2000; Moseley et al., 2013; Shergill et al., 2005; Whitford, 2019). Exposing voice-hearers to SOVD task and relating the corresponding SOVD microstate to the underlying behavior might thus either significantly support or challenge this prominent account.

A limitation of this work is that we do not have an assessment of SOVD prior to the removal of meningioma, that could be compared to the observed post-surgical SOVD. According to our assumption relating SOVD to personality alterations, we would observe diminished SOVD inversion effects before the resection. However, present findings still make a strong case for that claim, considering that out of 24 tested patients only this patient inverted self and other voices, and that only in this patient we observed significant personality alterations associated with the resection.

To conclude, here we report that major personality alterations which occurred following a resection of a large sagittal sinus meningioma can be evidenced in a SOVD task, thereby paving the way for development of novel biomarkers for the sense of self that could be used for pre- & post-surgical evaluation.

## Data Availability

All data produced in the present study are available upon reasonable request to the authors.

## ACKNOWLEDGMENTS

The authors thank the patient for her dedication and participation in the study, as well as the Swiss Foundation for Innovation and Training in Surgery (SFITS) in Geneva for hosting the experimental platform.

